# Kidney Allograft Recipients Diagnosed with Coronavirus Disease-2019: A Single Center Report

**DOI:** 10.1101/2020.04.30.20086462

**Authors:** Michelle Lubetzky, Meredith Aull, Rebecca Craig-Schapiro, Jun B. Lee, John R. Lee, Samuel Sultan, Jehona Marku-Podvorica, Laura Gingras, Rosy Priya Kodiyanplakkal, Choli Hartono, Stuart Saal, Thangamani Muthukumar, Sandip Kapur, Manikkam Suthanthiran, Darshana M. Dadhania

## Abstract

**Background:** Organ graft recipients receiving immunosuppressive therapy are likely to be at heightened risk for the Coronavirus Disease 2019 (Covid-19) and adverse outcomes including death. It is therefore important to characterize the clinical course and outcome of Covid-19 in this vulnerable population and identify therapeutic strategies that are safe.

**Methods:** We performed a retrospective chart review of 54 adult kidney transplant patients diagnosed with Covid-19 and all managed in New York State, the epicenter of Covid-19 pandemic. All 54 patients were evaluated by video visits, or phone interviews, and referred to our Fever Clinic or Emergency Room for respiratory illness symptoms consistent with Covid-19 and admitted if deemed necessary from March 13, 2020 to April 20, 2020. Characteristics of the patients were stratified by hospitalization status and disease severity. Clinical course including alterations in immunosuppressive therapy were retrieved from their electronic medical records. Primary outcomes included recovery from Covid-19 symptoms, acute kidney injury, graft failure, and case fatality rate.

**Results:** Of the 54 SARS-Cov-2 positive kidney transplant recipients, 39 with moderate to severe symptoms were admitted and 15 with mild symptoms were managed at home. Hospitalized patients compared to non-hospitalized patients were more likely to be male, of Hispanic ethnicity, and to have cardiovascular disease. At baseline, all but 2 were receiving tacrolimus, mycophenolate mofetil (MMF) and 32 were on a steroid free immunosuppression regimen. Tacrolimus dosage was reduced in 46% of hospitalized patients and maintained at baseline level in the non-hospitalized cohort. Mycophenolate mofetil (MMF) dosage was maintained at the baseline dosage in 11% of hospitalized patients and 64% of non-hospitalized patients and was stopped in 61% hospitalized patients and 0% in the non-hospitalized cohort. Azithromycin or doxycycline were prescribed at a similar rate among hospitalized and non-hospitalized patients (38% vs. 40%). In addition, 50% of hospitalized patients were treated for concurrent bacterial infections including pneumonia, urinary tract infections and sepsis. Hydroxychloroquine was prescribed in 79% of hospitalized patients and only one of 15 non-hospitalized patients. Acute kidney injury occurred in 51% of hospitalized patients. Patients with severe disease were more likely to have elevations in inflammatory biomarkers at presentation. At a median of 21 days follow up, 67% of patients have had their symptoms resolved or improved and 33% have persistent symptoms. Graft failure requiring hemodialysis occurred in 3 of 39 hospitalized patients (8%). Three of 39 (8%) hospitalized patients expired and none of the 15 non-hospitalized patients expired.

**Conclusions:** Clinical presentation of Covid-19 in kidney transplant recipients was similar to what has been described in the general population. The case fatality rate in our entire cohort of 54 kidney transplant recipients was reassuringly low and patients with mild symptomology could be successfully managed at home. Data from the our study suggest that a strategy of systematic screening and triage to outpatient or inpatient care, close monitoring, early management of concurrent bacterial infections and judicious use of immunosuppressive drugs rather than cessation is beneficial.

## Introduction

With the emergence of Coronavirus Disease 2019 (Covid-19) as a global pandemic, there are justifiable concerns regarding immunosuppressed organ graft recipients being at an increased risk for both contracting the virus and for adverse outcomes including death following the infection. Yet as we learn more about the virus and its mechanisms of injury, many questions remain in immunosuppressed solid organ transplant recipients including whether a reflex reduction in immunosuppressive therapy is an appropriate management strategy. This is in part because a large study screening existing FDA approved drugs for repurposing as antiviral therapeutics reported three different immunosuppressive agents widely used in transplant recipients-tacrolimus, mycophenolate, and sirolimus- as potential therapies for Covid-19 [1]. Moreover, in vitro studies have demonstrated that mycophenolic acid has activity against other coronaviruses, namely MERS-CoV [2]. It is possible therefore that immunosuppressive agents may actually confer protective effects against this particular virus, yet, the general early management strategy within the transplant community has been to reduce or withhold immunosuppression, particularly mycophenolate mofetil (MMF) in transplant recipients who are SARS-CoV-2 positive [3–6]. It is also unclear whether the cytokine release syndrome that can occur in response to SARS-CoV-2 infection and contribute to acute respiratory distress syndrome, is more likely or less likely to occur in patients who are on immunosuppressive therapy known to block transcriptional elements related to cytokine release. It is therefore critical to understand the clinical course and outcomes in the SARS-CoV-2 infected transplant recipients as compared to the general population so that treatment strategies can be optimized.

New York City is currently the Covid-19 epicenter of the world, with the prevalence rates of positive tests per New York zip code ranging from 27% to 78% in some of the most affected areas [7]. Since the first report of Covid-19 in 5 kidney graft recipients by Zhang et al. (3) several case reports of Covid-19 in kidney transplant recipients have been reported [3, 5, 6, 8–13].

At our institution, NewYork-Presbyterian/Weill Cornell Medicine (NYP/WCM), we developed a systematic approach for the evaluation of patients suspected to be infected with SARS-CoV-2 and a set of criteria for admission. In this report, we describe our center’s approach and the characteristics of our first 54 consecutive kidney allograft recipients confirmed to have Covid-19.

## Methods

We retrospectively studied adult (age>18 years) recipients of kidney allografts screened for mild or severe symptoms compatible with Covid-19 diagnosis at our outpatient transplant clinic, Weill Cornell Medicine (WCM) Fever Clinic or the NYP/WC Emergency Department (ED) from March 13, 2020 to April 20, 2020 and followed patients until April 25, 2020. A total of 71 outpatients reported symptoms suspicious for Covid-19; of those, 59 were referred to our center for evaluation; 54 patients tested positive for SARS-CoV-2, while 5 tested negative. The patients who tested negative for SARS-CoV-2 and patients who were not tested for the virus and monitored at home are not included in the analysis.

To identify variables that were associated with a more severe infection requiring hospitalization of kidney transplant recipients, we compared baseline and transplant parameters of kidney transplant patients who were hospitalized (n=39) to those managed in the outpatient setting (n=15). Our review was covered by our Weill Cornell Medicine Institutional Review Board approved protocol # 1207012637 entitled Utilizing a Transplant Database for Quality Assessment and Performance Improvement and Clinical Outcomes that collects data for our kidney transplant recipients for the purposes of quality improvement and clinical research. Procedures followed were in accordance with the ethical standards of the responsible committee on human experimentation (institutional and national) and with the Helsinki Declaration of 1975, as revised in 2013.

### Patient Screening and Triage

Our screening of kidney transplant recipients consisted of phone interviews and/or video visits. The initial approach for those with mild symptoms was to provide at-home supportive care. Only those patients where in-person evaluation was anticipated to impact their treatment were referred to NYP/WC Fever Clinic and/or ED. Those with fever >101 for >1 day and/or respiratory symptoms (including shortness of breath or dyspnea on exertion) were sent to NYP/WC Fever Clinic for evaluation. NYP/WC fever clinic is a Primary Care Clinic that was set up at the start of the Covid-19 pandemic to evaluate patients in the NYP/WCM community with symptoms suspicious for Covid-19 to make rapid diagnosis and importantly separate patients of suspected of Covid-19 from the general patient population seeking primary and subspecialty care. All patients presenting to the Fever Clinic had nasopharyngeal swab specimen collected by trained personnel and the sample was tested for the presence of SARS-CoV-2 via Polymerase Chain Reaction using one of the following tests: cobas® SARS-CoV-2, Xpert® Xpress SARS-CoV-2, or Panther Fusion® SARS-CoV-2 assay. Patients were sent to the ED if they met the following criteria: (1) hemodynamic instability, (2) volume depletion, (3) hypoxia with O2 saturation <94% on RA and/or (4) acute kidney injury (as defined by an increase of creatinine to ≥0.5mg/dL above baseline or a 30% increase from baseline).

### Anti-viral and anti-bacterial therapy

Admitted patients were evaluated by both a Transplant Physician and a Transplantation Infectious Disease specialist who guided anti-bacterial and/or antiviral therapies (including azithromycin, doxycycline and/or hydroxychloroquine). Additional antibiotics for bacterial pneumonia and/or sepsis were used at the discretion of the treating physicians.

Our standard protocol for hydroxychloroquine dosing included 600 mg BID x 2 doses, then 400 mg daily for 4 additional days. Patients were evaluated for inclusion in the remdesivir clinical trials if they met inclusion criteria for either the: (1) moderate severity trial - patients testing positive for SARS-CoV-2 who had symptoms of fever or cough, CXR with infiltrate, O2 sat >94% on room air and within 8 days of onset of symptoms- or (2) the high severity trial - O2 saturation <94% on room air within 4 days of a positive SARS-CoV-2 test.

### Data collection

Baseline demographics, co-morbidities, transplant details, immunosuppressive therapies, concomitant infections, treatment approaches, and clinical course were collected for all patients. Limited laboratory data were collected on those that were managed as outpatients using virtual video visits given the emphasis to minimize travel outside the home. Data were extracted from electronic medical record system.

### Data analysis

We performed detailed analysis of the 54 patients who tested positive for SARS-CoV-2 and compared the baseline and clinical parameters including patient and graft outcomes between those that required hospitalization (n=39) and those that did not (n=15). Of those that were hospitalized, we classified those requiring ventilator support or 100% non-rebreather mask as severe (n=12) and those that remained on room air or required oxygen by nasal cannula as moderate (n=27) and compared the admission presenting symptoms, laboratory test results, management.

## Results

### Characteristics of SARS-CoV-2 positive kidney allograft recipients

The first case of Covid-19 in a kidney transplant recipient was diagnosed at our center on March 13, 2020. Figure 1 shows the time course from March 8, to April 20, 2020 over which the 54 SARS-CoV-2 positive cases occurred. The peak of infections occurred during the week of April 5 to April 11, 2020, mirroring the peak of infections seen in New York City.

**Figure 1.**
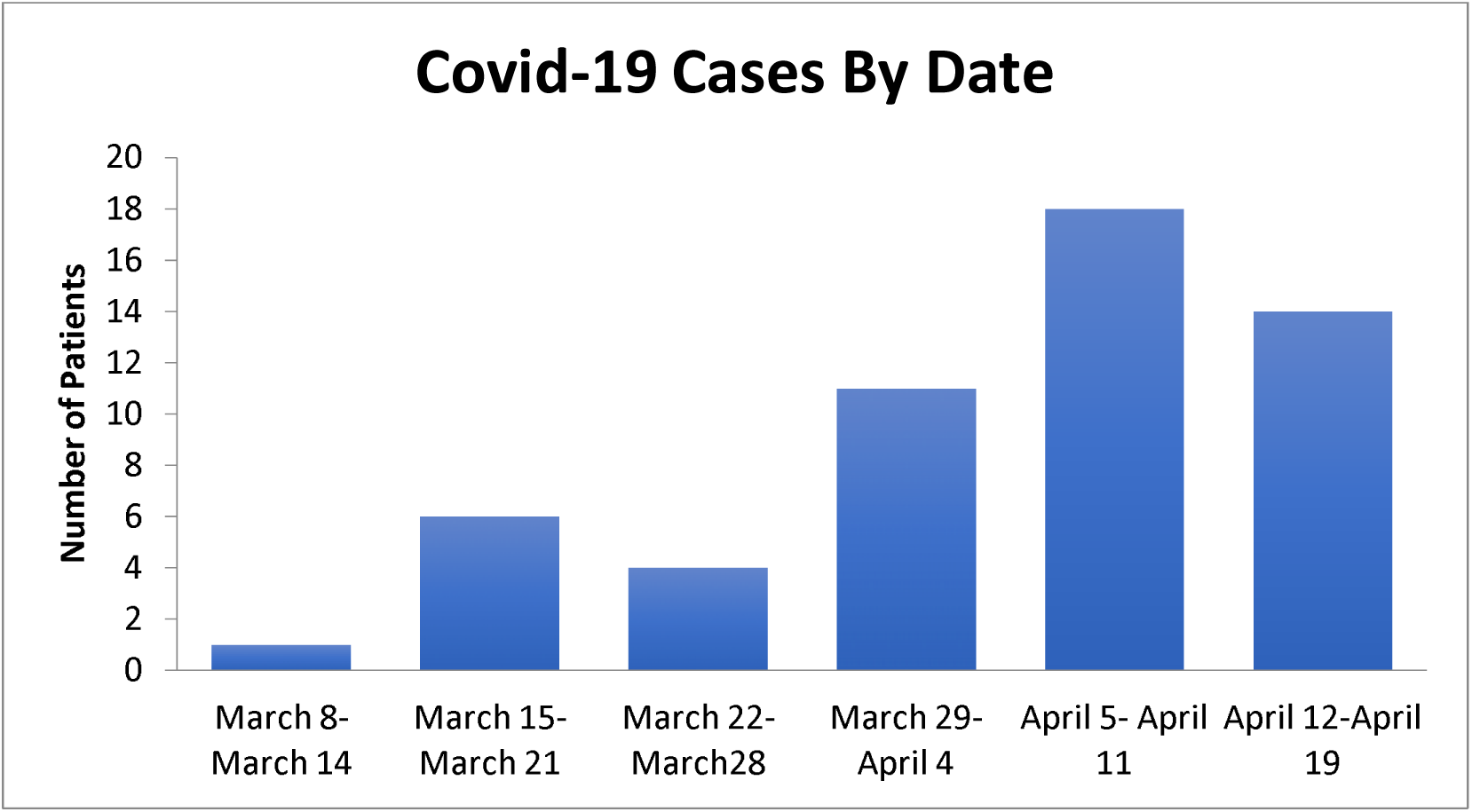
Weekly Cases of Kidney Transplant Patients with Covid-19.

Characteristics of all 54 SARS-Cov-2 positive kidney transplant recipients are listed in Table 1 and also separated into hospitalized cohort (n=39) and non-hospitalized cohort (n=15). Hospitalization was triggered by the severity of symptoms including shortness of breath, chest pain and/or persistent fever. The median age was 57 years in the entire cohort and 59 years in the hospitalized group and 55 years in the non-hospitalized group. The proportions of male (79% vs. 47%) and of Hispanic recipients (36% vs. 20%) were higher in the hospitalized cohort while the percentage of Caucasians (40% vs. 28%) was higher in the non-hospitalized group. ABO blood group type, BMI and smoking history were similar between those hospitalized and non-hospitalized. There was a greater prevalence of cardiovascular disease (41% vs. 20%) and pulmonary disease (18% vs. 7%) in the hospitalized patients compared to non-hospitalized patients. The prevalence of diabetes was similar between the two groups and the cause of ESRD was also similar between the two groups.

**Table 1.** Characteristics of Kidney Transplant Recipients with Covid-19.

|  | **Total** | **Hospitalized** | **Not Hospitalized** |
| --- | --- | --- | --- |
|  | N=54 | N=39 | N=15 |
| **Age, Years, median (range)** | 57 (29, 83) | 59 (29, 83) | 55 (31, 73) |
| **Sex, male, N (%)** | 38 (70) | 31 (79) | 7 (47) |
| **Race/Ethnicity, N (%)** |  |  |  |
| Caucasian/White | 17 (31) | 11 (28) | 6 (40) |
| Hispanic | 17(31) | 14(36) | 3 (20) |
| Black / African American | 13 (24) | 26(10) | 3 (20) |
| Asian | 6 (11) | 4 (10) | 2 (13) |
| Middle Eastern | 1 (2) | 0 (0) | 1 (7) |
| **ABO Blood Group Type, N (%)** |  |  |  |
| Type A | 15 (28) | 11 (28) | 4 (27) |
| Type B | 8 (15) | 6 (15) | 2 (13) |
| Type AB | 2 (3) | 2 (5) | 0 (0) |
| Type O | 29 (54) | 20 (51) | 9 (60) |
| **Smoking History, N (%)** | 12 (22) | 8 (21) | 4 (27) |
| **BMI, median, (IQR)** | 28 (18, 43) | 27 (18, 43) | 29 (18, 34) |
| **Co-Morbidities, N (%)** |  |  |  |
| Diabetes | 16 (30) | 12 (31) | 4 (27) |
| Cardiovascular Disease | 19 (35) | 16 (41) | 3 (20) |
| Stroke | 4 (7) | 3 (8) | 1 (7) |
| Pulmonary Disease | 8 (15) | 7 (18) | 1 (7) |
| Medications for Hypertension | 50 (93) | 37 (95) | 13 (93) |
| ACE inhibitor or ARB<sup>1</sup> | 19 (37) | 12 (32) | 7 (47) |
| **Cause of ESRD<sup>1</sup>, N (%)** |  |  |  |
| Hypertension | 11 (20) | 8 (21) | 3 (20) |
| Diabetes | 14 (26) | 11(28) | 3 (20) |
| Glomerulonephritis | 13 (24) | 11 (28) | 2 (13) |
| Lupus | 2 (4) | 1 (3) | 1 (7) |
| Polycystic Kidney Disease | 3 (6) | 1 (3) | 2 (13) |
| Other | 11 (20) | 7 (18) | 4 (27) |
| **Kidney Transplant Variables, N (%)** |  |  |  |
| **Transplant Type, living donor** | 37 (69) | 26 (67) | 11 (73) |
| **HLA A,B,DR Mismatch, median (range)** | 4 (0, 6) | 5, (0, 6) | 4 (2, 6) |
| **Donor Specific Antibodies at Transplant** | 15 (28) | 11 (28) | 4 (27) |
| **T cell depleting induction** | 39 (72) | 30 (77) | 9 (60) |
| **Steroid maintenance, N (%)** | 20 (37) | 14(35) | 6 (40) |
| **Prior acute rejection episode** | 4 (7) | 4 (10) | 0(0) |
| **Baseline serum creatinine (mg/dL), mean ± SD** | 1.52 ± 0.67 | 1.58 ± 0.74 | 1.34 ± 0.41 |
| **Viral infections, within past 3 months, N (%)** | 12 (22) | 9 (23) | 3 (20) |
| **Influenza** |  | 2 | 2 |
| **Coronavirus** |  | 1 | 1 |
| **RSV<sup>1</sup>** |  | 2 | 0 |
| **CMV<sup>1</sup>** |  | 2 | 0 |
| **BKV<sup>1</sup>** |  | 1 | 1 |
| **Other** |  | 2 | 0 |

<sup>1</sup>Abbreviations: ESRD (End Stage Renal Disease), ACE (Angiotensin-Converting Enzyme), ARB (Angiotensin Receptor Blocker), RSV (Respiratory Syncytial Virus), CMV (Cytomegalovirus), BKV (Polyomavirus BK)

### Kidney Transplant Related Variables

The type of organ donor (living vs. deceased donor), the number of HLA mismatches between the recipient and the donor, and the presence of preformed circulating DSA were similar in both groups. Induction therapy with T cell depleting antibodies and the proportion of patients on steroid-free maintenance immunosuppressive regimen (our standard-of-care protocol) were similar between both groups. The baseline mean ± SD serum creatinine was 1.58 ±0.74 mg/dL in the hospitalized cohort versus 1.34 ±0.41 mg/dL in the non-hospitalized cohort. Interestingly, approximately 22% of our patients had laboratory evidence of viral infection within 3 months of the diagnosis of SARS-Cov-2 with a similar incidence in hospitalized versus non-hospitalized group.

The median time since kidney transplantation to SARS-Cov-2 diagnosis was 4.7 years (range of 0.3 to 35 years) (Table 2). Both hospitalized and non-hospitalized cohorts included patients within 1 year of transplantation at a similar rate (18% vs. 13%). Tacrolimus is the calcineurin inhibitor of choice in our kidney transplant recipients and the tacrolimus dose was higher in the hospitalized recipients compared to non-hospitalized recipients (4.8±2.67 mg/day vs. 4.12±2.48 mg/day) whereas the total dose per day of mycophenolate mofetil (MMF) was lower in hospitalized group compared to not-hospitalized patients (1.18 ± 0.48 grams per day vs 1.30 ± 0.52 grams per day). Thirty-seven of 54 patients were on steroid free maintenance protocol. Two of the 54 patients were on calcineurin inhibitor free protocol, one on everolimus and one on belatacept. Two of 54 kidney transplant recipients diagnosed with Covid-19 did not receive MMF as maintenance therapy and were instead on mTOR inhibitor rapamycin (n=2).

**Table 2.** Kidney Transplant Recipients with Covid-19: Presenting Symptoms.

|  | **Total** | **Hospitalized** | **Not Hospitalized** |
| --- | --- | --- | --- |
|  | N=54 | N=39 | N=15 |
| **Covid-19 Diagnosis** |  |  |  |
| **Post-Transplant Years, median (range)** | 4.7 (0.3, 35) | 4.7 (0.3, 14.4) | 4.6 (0.5, 35.3) |
| **Days from symptoms to diagnosis, mean (± SD)** | 8.2 ± 6.0 | 8.5 ± 6.6 | 7.6 ± 4.5 |
| **Covid-19 Exposure** | 25 (46) | 16 (41) | 9 (60) |
| **Home** | 14 (26) | 10 (26) | 4 (27) |
| **Nursing home / hospital** | 4 (7) | 3 (8) | 1 (7) |
| **Work** | 6 (11) | 2 (5) | 4 (27) |
| **No known** | 30 (56) | 24 (62) | 6 (40) |
| **Immunosuppression at diagnosis, N (%)** |  |  |  |
| **Tacrolimus dose (mg/day) at diagnosis<sup>1</sup>** | 4.65 ± 2.62 | 4.8 ± 2.67 | 4.12 ± 2.48 |
| **Mycophenolate mofetil dose (g/day) at diagnosis<sup>2</sup>** | 1.21 ± 0.49 | 1.18 ± 0.48 | 1.30 ± 0.52 |
| **Steroid maintenance** | 22 (41) | 16 (41) | 6 (40) |
| **Presenting Symptoms, N (%)** |  |  |  |
| **Fever** | 40 (74) | 30 (77) | 10 (67) |
| **Cough / upper respiratory symptoms** | 32 (59) | 22 (56) | 10 (67) |
| **Shortness of breath** | 28 (52) | 21 (54) | 7 (47) |
| **Fatigue / myalgia** | 23 (43) | 17 (44) | 6 (40) |
| **Diarrhea** | 21 (39) | 15 (38) | 6 (40) |
| **Nausea / vomiting** | 5 (9) | 4 (10) | 1 (7) |
| **Confusion** | 6 (11) | 6 (15) | 0 (0) |
| **Viral Pneumonia Diagnosis<sup>3</sup>, N (%)** | 36/42 (86) | 32/37 (86) | 4/5 (80) |
| **unilateral presentation** |  | 4/37 (11) | 1/5 (20) |
| **bilateral presentation** |  | 28/37 (76) | 3/5 (60) |

<sup>1</sup>based on 52 patients on tacrolimus, 1 on everolimus, 1 on belatacept<sup>2</sup>data available on 38 in hospitalized and 4 in the not hospitalized group<sup>3</sup>data available on 37 of the hospitalized and 5 in the not hospitalized group

### Covid-19 Illness Presentation

The most common presenting symptom was fever (74%), followed by cough (59%), shortness. of breath (52%), myalgias/fatigue (43%), and gastrointestinal symptoms of diarrhea (39%) and nausea/vomiting (9%) (Table 2). Of the entire cohort of 54 patients, 42 (78%) underwent a chest x-ray as part of their evaluation and the predominant finding was bilateral patchy airspace opacities in the lower lung fiends in both groups. Forty-eight percent of our patients overall and 62% of those hospitalized were diagnosed with a concurrent bacterial infection and were treated with additional antibiotic therapy.

### Management of Covid-19 in Kidney Transplant Recipients

We elected to continue immunosuppressive drug therapy but at a reduced level in most of our patients. Tacrolimus dosage was adjusted downwards so that the tacrolimus trough levels were 4-6ng/ml. Tacrolimus dosage was reduced from baseline in 46% of hospitalized patients and no adjustments were made in patients managed at home. MMF was continued at baseline dosage in 9 of 14 non-hospitalized patients, reduced by 50% in the remaining 5 patients (Table 3). In the hospitalized patients, a more aggressive reduction in MMF dosage was undertaken with MMF being withheld in 24 patients (61%), a 50% dose reduction in 10 patients (26%) and no reduction in the remaining 4 patients (11%).

**Table 3.** Medical Management of Covid-19 in Kidney Transplant Recipients.

|  | **Total** | **Hospitalized** | **Not Hospitalized** |
| --- | --- | --- | --- |
|  | N=54 | N=39 | N=15 |
| **Adjustment in Tacrolimus dose<sup>1</sup>, N (%)** |  |  |  |
| Reduction from baseline dose | 17(33) | 17(46) | 0 (0) |
| No adjustment from baseline | 35 (67) | 20 (54) | 15 (100) |
| Held | 0 (0) | 0 (0) | 0 (0) |
| **Adjustment in Mycophenolate mofetil dose<sup>2</sup>, N (%)** |  |  |  |
| No reduction / less than 50% reduction | 13 (28) | 4 (11) | 9 (64) |
| 50% reduction | 15 (28) | 10 (26) | 5 (33) |
| Discontinued drug | 24 (44) | 24 (61) | 0 (0) |
| **Adjustment in steroid dose, N (%)** |  |  |  |
| Continued maintenance steroid<sup>3</sup> | 22 (41) | 16 (41) | 6(40) |
| Remained steroid free<sup>4</sup> | 29 (54) | 20 (51) | 9 (60) |
| Additional steroid therapy | 5 (9) | 5 (13) | 0 (0) |
| **ACE inhibitor / ARB discontinued<sup>5</sup>** | 11 (58) | 10 (83) | 1 (14) |
| **Antibiotics, N (%)** | 21 (39) | 15 (38) | 6 (40) |
| Azithromycin | 12 (22) | 7 (18) | 5 (33) |
| Doxycycline | 8 (17) | 8 (21) | 1 (7) |
| **Experimental Therapies, N (%)** |  |  |  |
| Hydroxychloroquine | 32 (62) | 31 (79) | 1 (7) |
| Remdesivir | 2 (4) | 2 (5) | 0 (0) |
| IL-6 receptor inhibitor | 2 (4) | 2 (5) | 0 (0) |
| Convalescent Plasma | 1 (2) | 1 (3) | 0 (0) |
| **Management of bacterial infection at diagnosis<sup>6</sup>, N (%)** | 25 (48) | 23 (62) | 2 (13) |
| Treated for Urinary tract infection | 6 (11) | 4 (10) | 2(13) |
| Treated for Presumed bacterial pneumonia | 15 (28) | 15(38) | 0(0) |
| Treated for SIRS/sepsis<sup>7</sup> | 4 (7) | 4 (10) | 0(0) |

<sup>1</sup>Based on 52 patients on tacrolimus; 2 calcineurin inhibitor free, 1on everolimus, 1on belatacept
<sup>2</sup>Based on 52 patients on mycophenolate mofetil, 2 were on sirolimus
<sup>3</sup>Based on 22 patients on maintenance steroids
<sup>4</sup>Based on 32 patients on steroid free immunosuppression
<sup>5</sup>Based on 19 patients on ACE/ARB
<sup>6</sup>Based on data from 52 patients, 37 hospitalized patients
<sup>7</sup>Abbreviations, SIRS, systemic inflammatory response syndrome

Prednisone was continued in the 22 patients on a steroid maintenance regimen and the 29 of 32 patients on a steroid free protocol remained steroid free. Five patients received additional steroids during hospitalization. Prednisone was not withheld in any patient who was receiving steroids on admission.

For Covid-19, those requiring supplemental oxygen were initiated on hydroxychloroquine therapy with 79% of hospitalized patients receiving hydroxychloroquine. Among those that did not require hospital admission, only 1 outpatient received hydroxychloroquine while 1 was already on hydroxychloroquine for systemic lupus erythematosus. In addition, equal number of hospitalized and non-hospitalized patients received empiric treatment with azithromycin or doxycycline (38% and 40%) for symptoms of respiratory illness (Table 3).

Among those with severe illness, 9 received additional therapies; 5 of 9 received additional corticosteroids, 2 received treatment with remdesivir as part of a study protocol, 1 received treatment with Interleukin 6 (IL-6) receptor antagonist and 1 received both IL-6 receptor antagonist and convalescent plasma (Table 3).

### Hospital Course and Outcomes

Among those hospitalized, 12 were classified as having severe illness based on the need for either ventilator support or 100% non-rebreather mask. Those classified as having severe disease, had increased levels of C-reactive protein, procalcitonin, d-dimer and IL-6.

Detailed information including vitals and laboratory test results are shown in Table 4 stratified by severity of illness. On presentation, 69% of patients required supplemental O2. Most patients were not hypotensive or febrile upon admission and only 5 patients (14%), all in the severe illness category required pressor support during their hospital course and 9 required intubation. No patients required extracorporeal membrane oxygenation therapy (ECMO). Our patients all were relatively lymphopenic upon admission. Although not all data was available on admission to hospital, patients who had more severe illness were more likely to have elevated d-dimer, C-reactive protein and procalcitonin levels (Table 4).

**Table 4.** Kidney Transplant Recipients with Covid-19: Admission Vitals and Laboratory Values in Hospitalized Patients.

|  | All Hospitalized<br>N=39 | Severe Illness<br>N=12 | Moderate Illness<br>N=27 |
| --- | --- | --- | --- |
| **Admission Vitals** |  |  |  |
| Temperature (C) | 37.7 (37.1, 38.2) | 37.5 (37, 37.8) | 37.7 (37.1, 38.4) |
| Oxygen Saturation, median (IQ range) | 93 (89, 96)<br>94 (86, 98) | 90 (87, 93)<br>93 (88, 96) | 95 (91, 98)<br>96 (93, 98) |
| Respiratory Rate | 20 (18, 23) | 23 (19, 29) | 18 (18, 20) |
| Heart Rate | 94 (81, 107) | 93 (80, 106) | 94 (83, 107) |
| Systolic Blood Pressure (mmHg) | 128 (114, 141) | 124 (112, 154) | 129 (114, 139) |
| **Laboratory Values** |  |  |  |
| Creatinine (mg/dL), mean ± SD | 2.6 ± 2.3 | 3.1 ± 3.2 | 2.5 ± 1.8 |
| Acute kidney injury<sup>1</sup>, N (%) | 20(51) | 7 (58) | 13 (48) |
| White Blood Cell Count<sup>2</sup><br>($\times 10^3/\mu\text{L}$) | 5.7 (3.6, 8) | 5.5 (4.1, 9.9) | 5.9(2.5, 7.5) |
| % Lymphocytes<sup>2</sup> |  |  |  |
| Absolute lymphocyte count<sup>2</sup><br>($\times 10^3/\mu\text{L}$) | 11.4 (6.6, 17.1)<br>0.6 (0.3, 1.0) | 10.8 (3.9, 16.9)<br>0.6 (0.3, 0.9) | 11.8 (6.9, 17.4)<br>0.6 (0.3, 1.0) |
| Albumin (g/dL)<sup>2</sup> |  |  |  |
| C-reactive protein (mg/dL)<sup>2</sup> | 3.1 (2.7, 3.6) | 3.0 (2.6, 3.6) | 3.2 (2.8, 3.6) |
| Procalcitonin (ng/mL)<sup>2</sup> | 11.4 (5.3, 30.2) | 16.5 (8.6, 22.1) | 10.1 (5.2, 35) |
| D-Dimer (ng/mL)<sup>2</sup> | 0.3 (0.1, 0.6) | 0.6 (0.3, 1) | 0.2 (0.1, 0.4) |
| Ferritin (ng/mL)<sup>2</sup> | 394 (278, 589) | 506 (306, 755) | 365 (252, 499) |
| Interleukin 6 (pg/mL)<sup>2</sup> | 1498 (383, 2646)<br>8 (4.5, 92) | 1152 (616, 2440)<br>170 (14, 268) | 1882 (307, 2646)<br>4.5 (3, 9) |

<sup>1</sup>Acute kidney injury as defined by absolute creatinine increase of $\geq 0.5\text{mg/dL}$ or 30% increase from baseline creatinine
<sup>2</sup>Data based on the following numbers of patients, creatinine, white blood cell and lymphocyte count n=37, procalcitonin n=35, albumin n=34, C-reactive protein n=29, ferritin n=28, d-dimer n=26, Interleukin-6 n=9.

Patients were followed for a median of 21 days (range 5, 43) from symptom onset, with a median of 21 days for those hospitalized and 19 days for those not hospitalized. The median number of days of hospitalization was 8 days with a range of 5 to 22 days at last follow-up; 42% had greater than 10 hospital days. We classified the patients as having Covid-19 symptoms ‘resolved’ if they were at home and no longer symptomatic, ‘improved’ if some of their symptoms were still present but overall feeling better and ‘not improved’ if they remained symptomatic or hospitalized. Figure 2A shows the percentage of patients with the outcome of Covid-19 symptoms in those hospitalized as well as those not hospitalized, as of April 25, 2020.

**Figure 2A.**
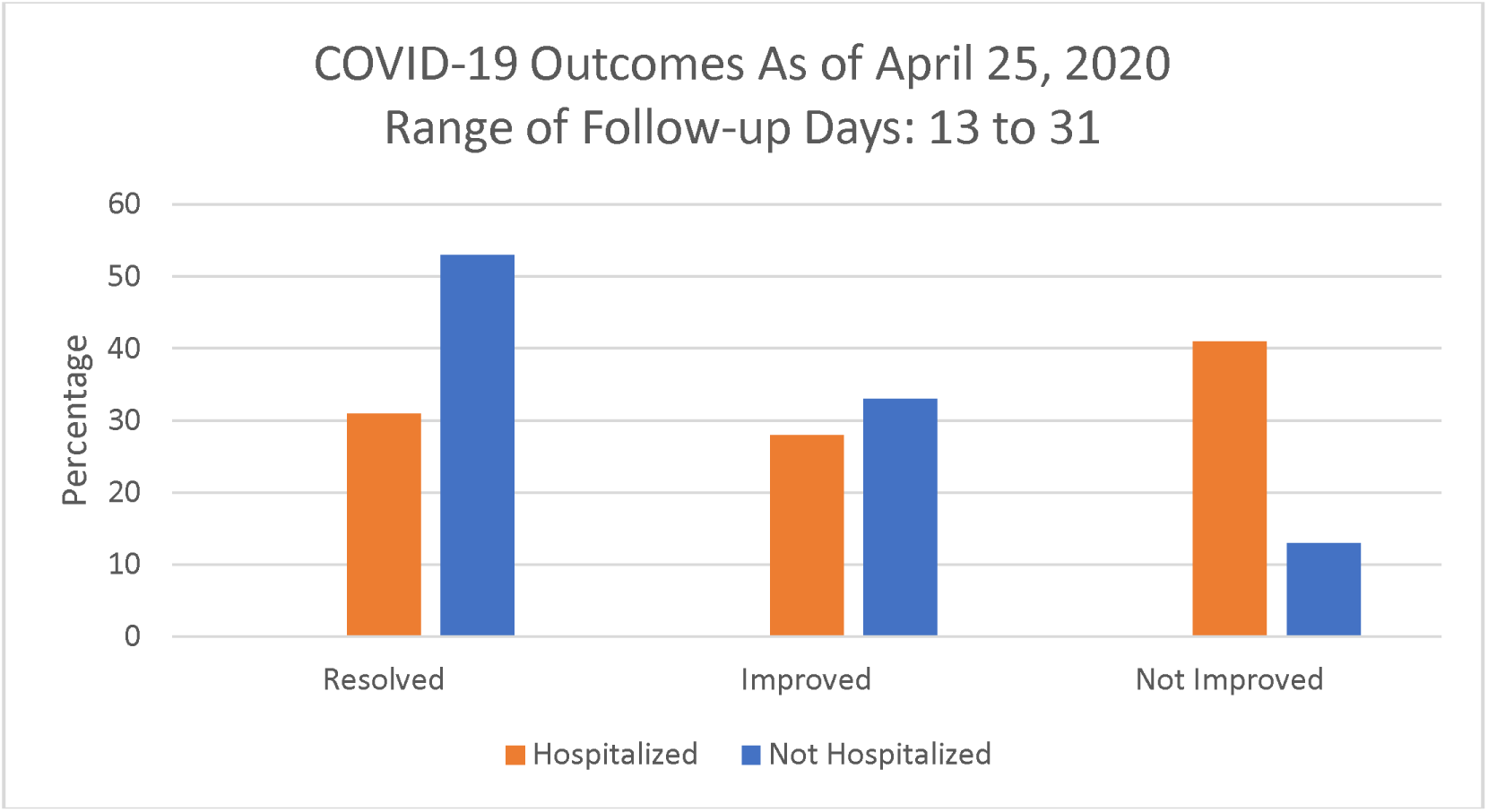
Covid-19 Symptoms.

Twenty patients (51%) who were admitted to the hospital developed acute kidney injury during the course of their illness (Table 5). At last follow-up, 35% had resolution of their AKI, 30% had partial resolution and the remaining 35% did not have resolution of their AKI (Figure 2B). Five of 39 hospitalized patients (13%) required initiation of renal replacement therapy and in 7 of 39 (18%) the estimated GFR was less than 20 ml/minute (Figure 2B).

**Table 5.** Outcomes in Kidney Transplant Recipients with Covid-19.

|  | **Total** | **Hospitalized** | **Not Hospitalized** |
| --- | --- | --- | --- |
|  | N=54 | N=34 | N=15 |
| **Acute Kidney Injury<sup>1</sup>** | 21(39) | 20 (51) | 1(7) |
| **Outcomes of AKI** |  |  |  |
| **Resolved<sup>2</sup>** | 7 (35) | 7 (37) | 0 (0) |
| **Partially resolved<sup>2</sup>** | 6 (30) | 5 (26) | 1 (100) |
| **Not resolved** | 7(35) | 7 (37) | 0 (0) |
| **Graft outcome** |  |  |  |
| **eGFR<20cc/min** | 8 (15) | 8 (21) | 0 (0) |
| **Remains Hospitalized** | 11 (20) | 11 (32) | 0 (0) |
| **Patient Death** | 3 (6) | 3 (8) | 0 (0) |

<sup>1</sup> Acute kidney injury as defined by absolute creatinine increase of $\geq 0.5$ mg/dL or 30% increase from baseline creatinine
<sup>2</sup> Resolved AKI as defined by a return to within 15% of baseline creatinine at last follow up and partially resolved as defined by an improvement of serum creatinine that is not within 15% of baseline

**Figure 2B.**
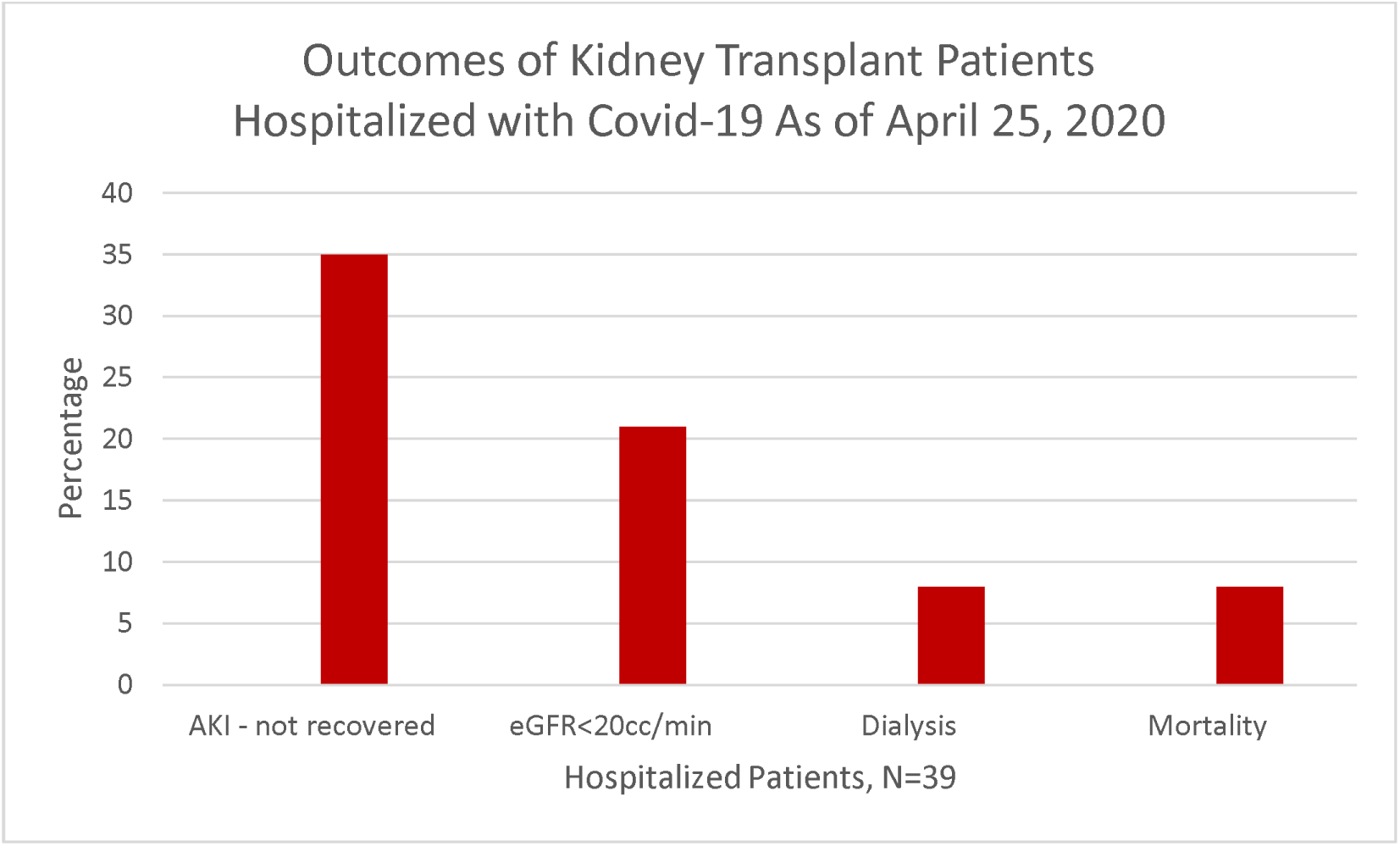
Complications.

Of the 39 that were hospitalized, 3 of 39 hospitalized patients died (7.7%), 25 (64%) were discharged, and 11 (32%) remain hospitalized. All 15 non-hospitalized patients are alive as of April 25, 2020. The case fatality rate among all 54 Covid-19 patients was 5.5% and among the 39 hospitalized patients was 7.7% (Table 5 and Figure 2B).

## Discussion

Despite initial fears that transplant patients may be amongst those at the highest risk of adverse outcomes during the Covid-19 pandemic, our initial results comprised of 54 kidney allograft recipients diagnosed with Covid-19 demonstrate that a coordinated outpatient and inpatient effort can effectively manage kidney transplant recipients with Covid-19. With a median follow up of 21 days, our total overall mortality was 5.5% and hospitalized mortality rate was 8%. Unique aspects of our management include (1) careful adjustment of immunosuppressive therapies; (2) aggressive evaluation and management of secondary bacterial infections, and (3) judicious and monitored use of unproven therapies such as hydroxychloroquine. By systematically evaluating patients via telemedicine applications and coordinating our outpatient laboratory procedures, many patients were able to be successfully managed in the outpatient setting and at home. Patients with more severe symptoms were sent directly to the Covid-19 triage area of the NYP/WCM Emergency Department or to the NYP/WC Fever Clinic, thus helping to minimize the risk of infecting other transplant patients who were coming to the transplant clinic for routine follow up. Once hospitalized at NYP-WCM, an interdisciplinary team that included transplant nephrologists, transplant infectious disease specialists and transplant surgeons followed the patients. With this approach we were able to obtain a relatively low case fatality rate of 5.5% in the entire cohort of 54 kidney allograft recipients.

Risk factors for more severe disease in our transplant population mirrored those reported in the general Covid-19 population[14]. In our cohort, patients who were hospitalized had more comorbidities, specifically cardiovascular disease, and were more likely to have more severe disease and require admission to the hospital. Additionally, 14 out of the 39 kidney transplant recipients who were hospitalized were over 65. Patients with more severe disease also were found to have elevations of ferritin, d-dimer, procalcitonin, IL-6, and C-reactive protein which are similar to what has been reported in other cohorts [15].

One of the most pressing questions currently is how to manage immunosuppression in transplant recipients diagnosed with Covid-19. The general consensus in the transplant community thus far appears to be to decrease or withhold antimetabolites like mycophenolic acid [3, 6, 10, 12, 16]. There is less agreement about the optimal management strategy for calcineurin inhibitors, however. There was heterogeneity in the choice of immunosuppression adjustment even within our own cohort. Although the optimal management is not known, a possible strategy is to reduce MMF to 50% of baseline dose and target tacrolimus level at 4-6ng/dL for those with mild-moderate infection, and to more aggressively reduce or withhold MMF in the setting of more severe Covid-19 infection. Thus, the majority of our patients we managed in the outpatient setting had no change in their immunosuppression regimens. Upon admission to the hospital, a reduction in their MMF dosing was undertaken, and in those with severe illness, MMF was held. We also noticed that some patients required tacrolimus dose adjustments due to supratherapeutic levels observed at admission. Despite lowering and in some cases withholding of MMF, there were no confirmed cases of acute rejection in our study cohort. However, due to lack of kidney transplant biopsies in the setting of AKI, the true incidence of acute rejection in our study is not known. Upon discharge from the hospital, immunosuppressive medications that were held were re-initiated and doses were slowly increased to baseline levels.

Despite the lack of clear evidence regarding its efficacy, the majority of our hospitalized patients received hydroxychloroquine. Hydroxychloroquine, in combination with azithromycin, has been associated with QT prolongation and potential adverse effects, such as cardiac arrhythmias [17, 18]. In our cohort, the decision to treat with azithromycin or doxycycline was not linked to the use of hydroxychloroquine but related to respiratory symptoms. Among our patients only one patient developed prolonged QTc causing the duration of therapy to be shortened and two patients experienced new onset atrial fibrillation. Further complicating the use of hydroxychloroquine is that results regarding efficacy in treating SARS-Cov-2 has been conflicting. A non-randomized twenty patient study showed that hydroxychloroquine reduced viral load, but failed to correlate results with clinical outcomes, provided the basis for many centers’ adoption of this experimental therapy [19]. A subsequent, larger non-randomized study, failed to show reduction in risk of mechanical ventilation, and demonstrated increased mortality with treatment [20]. It remains to be seen whether hydroxychloroquine use in large, randomized, placebo-controlled trials will yield beneficial results. No definite conclusions can be drawn from our study regarding hydroxychloroquine efficacy in transplant patients, although majority of our outpatients did not receive hydroxychloroquine and 86% had improvement or resolution of symptoms. In light of the excellent outcome observed in our patients managed in the outpatient setting, we would recommend against routine use of hydroxychloroquine in kidney transplant recipients especially in the outpatient setting where fatal cardiac rhythm abnormalities may go undetected.

Only two of our patients were enrolled in remdesivir studies, one of which did not require intubation and was discharged from the hospital and the other remains inpatient but is no longer in the ICU. While initial data on the compassionate use of remdesivir seems promising, follow up data and specific outcomes in solid organ transplant patients enrolled in ongoing trials is yet to be reported [21]. Two of our patients received IL-6 receptor antagonists. Both are still hospitalized and it unclear whether this strategy is useful in kidney transplant recipients.

There is emerging data that immunosuppressive agents used in kidney transplant recipients may provide yet unappreciated benefit against this particular virus. In vitro studies have demonstrated a potential role for immunosuppressive agents, including tacrolimus, sirolimus and MMF as viral targets [1, 2, 22, 23]. Additionally, one of the most serious complications of SARS-CoV-2 infection is the cytokine storm contributing to ARDS. Some immunomodulatory agents such as interleukin-6 and interleukin-1 antagonists, IVIG and steroids have been suggested to have a role in combatting this inflammatory response [24], Calcineurin inhibitors may be potentially beneficial in this setting because of their ability to reduce cytokine production via inhibition of nuclear localization of the transcription factor NFAT [25]. It is worth noting that all patients in our study were continued on tacrolimus albeit at reduced dosage and with due consideration of tacrolimus trough levels. Altogether, our data suggest immunosuppression reduction rather than cessation may be a reasonable approach especially when viewed through the lens of their ability to inhibit cytokine production and their potential anti-viral activities.

In our series, we observed a higher rate of acute kidney injury than reported in the general Covid-19 population, which has ranged from 3-9% in early studies to 15% in more recent studies [26]. While 16% of all Covid-19 related hospitalizations at our center had acute kidney injury (7), 51% of our hospitalized transplant patients experienced AKI. AKI in Covid-19 is thought to be a sequela from sepsis and cytokine storm syndrome; however, SARS-CoV-2 has been found in the kidneys and urine of COVID-19 patients suggesting there may be a direct mechanism of renal injury as well [26, 27]. It is hypothesized that the virus can directly infect renal tubules, resulting in acute tubular damage, and induces CD68+ macrophage and complement C5b-9 mediated damage [27]. Zhang and colleagues proposed a genetic mechanism via the expression of human ACE2 in the kidney and kidney specific expression quantitative train loci of potential direct viral targets in the kidney. There are several potential reasons why the transplant population may be more predisposed to acute renal injury during Covid-19 infection. First, many kidney transplant recipients have baseline stage 2 or 3 chronic kidney disease which may have contributed to AKI, especially in the setting of an acute viral illness. Elevated tacrolimus trough levels were observed in several of our patients at admission. It has been reported that the bioavailability of tacrolimus is increased due to reduced gut transit time with diarrhea (28). The high trough levels we observed may be a consequence of diarrhea and other gastrointestinal disturbances in our study cohort. Despite the high rates of AKI in our cohort, most patients (60%) either had full or partial renal recovery at most recent follow up. Thus, while Covid-19 primarily manifests as a respiratory disease, there is clearly an important kidney component, and the long-term prognosis with regards to kidney function warrants further study.

Eleven of the 39 hospitalized patients remain hospitalized at this time. The 8% in-hospital case fatality rate observed in our study is lower than the 28% mortality rate reported from a New York City transplant center and not dissimilar to the 10.2% mortality rate of non-transplant Covid-19 patients hospitalized at our center [7, 14]. Given that our study is comprised of only 54 kidney allograft recipients and with the follow up is limited, it is unclear whether our positive experience will translate to other centers or hold up with a longer follow up period.

Limitations of this study that must be considered include the small sample size. Additionally, due to limitations in Covid-19 testing capabilities in New York, we were unable to test all patients with symptoms and thus the number of infected transplant patients is likely much higher. Since remdesivir or IL-6 receptor antagonist was not given in a systematic fashion in our study, we will have to wait for the results from the controlled trials to understand their effectiveness. Yet it is important to note that many transplant patients may not be eligible for such studies which is why careful review of single center data will be an integral piece to understand how best to treat SARS-Cov-2 infections in kidney transplant recipients.

In sum, using a coordinated and multidisciplinary approach, the patients with mild symptoms, were successfully managed as outpatients with close monitoring for symptom progression and with minimal reductions in immunosuppressive agents. For hospitalized patients, careful evaluation and judicious reduction in immunosuppressive drugs and prompt treatment of secondary bacterial infections, were associated with a low case fatality rate. We did however observe significant AKI in a substantial percentage of hospitalized patients. A therapeutic strategy of clinical severity dependent reduction rather than complete withdrawal of immunosuppressive drug therapy appears reasonable for kidney allograft recipients diagnosed with Covid-19.

## Data Availability

The data included is clinical data on patients and could be further de-identified if needed but is not currently available

## Acknowledgements

We would like to thank all members of the Division of Nephrology and Transplant Surgery for their excellent care of our transplant patients and the WCM Fever Clinic for accommodating prompt evaluation of SARS-Cov-2 infection in our patient population. Early findings of 12 out of the 54 patients included in this series were included in a study by Pereira et al.[13].

## Conflicts of Interest

J.R.L. receives research funding support from BioFire Diagnostics, LLC. The other authors of this manuscript do not report any competing interests as it relates to these data and the material presented in this manuscript.

## Author’s Contributions

All authors participated in the care of these patients, provided clinical management details and reviewed the manuscript for accuracy, interpretation of the data and completeness. ML, MA, RS, JM and DMD participated in the data collection and analysis of the data.

## Funding

The study was supported by internal divisional funds.

